# Modelling the potential use of pre-exposure prophylaxis to reduce nosocomial SARS-CoV-2 transmission

**DOI:** 10.1101/2024.12.27.24319372

**Authors:** Lauren Stewart, Stephanie Evans, Teresa Brevini, Fotios Sampaziotis, Christopher J. R. Illingworth

## Abstract

The nosocomial transmission of respiratory pathogens is an ongoing healthcare challenge, with consequences for the health of vulnerable individuals. Outbreaks in hospitals can require the closure of bays or entire wards, reducing hospital capacity and having a financial impact upon healthcare providers. Here we evaluate a novel strategy of pre-exposure prophylaxis as a means to reduce the nosocomial transmission of SARS-CoV-2. We model the effect of ursodeoxycholic acid (UDCA) upon levels of ACE2 expression, SARS-CoV-2 viral entry, and ultimately the probability of an infection. We then implement this model within simulations describing the spread of SARS-CoV-2 infections within a hospital context, simulating an intervention in which UDCA is given to patients on a ward for 10 days following the detection of a case of SARS-CoV-2 on that ward. Under default model parameters we infer a potential 16.5% reduction (95% C. I. 14% - 20%) in the nosocomial transmission of SARS-CoV-2 to patients, with increased importation of cases into the hospital increasing the effectiveness of the intervention. Our study provides preliminary evidence of the value of pre-exposure prophylaxis with UDCA as a strategy to reduce nosocomial SARS-CoV-2 transmission.

## Introduction

The SARS-CoV-2 virus has had a major impact upon human health^1,2^. As such, a key priority has been the identification of public health interventions which either prevent or reduce the impact of infection.

Vaccines have been shown to reduce the chances of infection leading to hospitalisation or death^3^, albeit that the immunity gained from vaccination wanes over time^4^, while virus evolution creates a need to continually re-evaluate vaccine effectiveness^5^. Other lines of defence against SARS-CoV-2 include antiviral drugs, such as pavloxid^6^, remdesivir^7^ and molnupiravir^8^. The US Food and Drug Administration has issued an emergency use authorisation for a monoclonal antibody therapy for pre-exposure prophylaxis^9^. These interventions have their own limitations, with for example a five-day course of molnupiravir being associated with a lower SARS-CoV-2 viral load five days after treatment, but a higher viral load after 14 days^10^. Non-pharmaceutical interventions, such as handwashing, masking, and social distancing, have helped to prevent infection^11^.

Hospitals and care homes have been the focus of particular attention in the monitoring and prevention of SARS-CoV-2 transmission. Even as the consequences of COVID have attenuated for the general population^12^, the concentration of potentially vulnerable individuals in these environments creates an enhanced need for action. One study early in the pandemic estimated that close to 15% of SARS-CoV-2 cases in hospital were the result of hospital-acquired infection^13^. Accordingly, studies have evaluated the use of mask-wearing by health care workers and patients in hospitals^14,15^. The installation of air cleaning units on a hospital ward was shown to reduce the concentration of airborne particulates of a size commensurate with airborne viral transmission^16^. Air cleaning has been associated with a 22% reduction in nosocomial transmission (95% CI 47% to −14%)^17^.

A mathematical study identified pre-exposure prophylaxis (PrEP) as being potentially the most effective complement to vaccination in preventing SARS-CoV-2 infection^18^. Mathematical modelling provides a valuable insight into potential therapeutic approaches in advance of committing financial and clinical resources to a real-world trial. However, PrEP approaches to combatting SARS-CoV-2 are not simply theoretical constructs. A Mendelian randomisation study of potentially druggable proteins identified the genes ACE2 and IFNAR2 as priority targets for intervention^19^. The former is a cell receptor which critically must be bound by SARS-CoV-2 Spike protein prior to cellular entry^20^, while the latter participates in the host innate immune response^21^ against the virus. ACE2 expression has been associated with more severe clinical outcomes of SARS-CoV-2 infection^22^.

The discovery that the farnesoid X receptor (FXR) regulates ACE2 expression provided a novel approach to controlling SARS-CoV-2 infection^23^. Ursodeoxycholic acid (UDCA) is a well-tolerated and off-patent drug which downregulates FXR, leading to a reduction in ACE2 expression. A cohort study identified a significant reduction in the odds of developing SARS-CoV-2 infection among patients with cirrhosis who were treated with UDCA^24^, finding also a reduced risk of disease severity among treated individuals. Although another study found no significant reduction in the risk of hospitalisation for COVID among treated individuals^25^, the basic result has been replicated in independent studies^26,27^, with high adherence to UDCA treatment^28^, and an increased dose of UDCA^29^ being associated with reduced rates of infection. A contrary result was reported in a smaller cohort of patients^30^.

While clinical studies have compared individuals treated with UDCA to those not receiving the drug, there may exist a broader scope to use the drug in a responsive fashion, for example as pre-exposure prophylaxis given an anticipated period of greater risk of SARS-CoV-2 infection.

In this study we use a mathematical model to evaluate the potential for UDCA to reduce nosocomial transmission. Our model encompasses the multiple scales on which the drug affects transmission, including the time-dependent effect of UDCA upon ACE2 expression, the relationship between ACE2 expression and SARS-CoV-2 viral entry into cells, and the distribution of levels of exposure occurring during a potential transmission event. Briefly, our model describes the effect of UDCA in terms of a change in the distribution of effective levels of viral exposure. SARS-CoV-2 exposures vary according to a range of individual and environmental factors^31^, such as the distance from an infected host. We characterise exposure in units of the number of viruses expected to initiate infection, using published data describing transmission bottlenecks^32^ and secondary attack rates^33^ for SARS-CoV-2 to derive an exposure distribution (Figure 1A). Treatment with UDCA reduces ACE2 expression (Figure 1B), and therefore the proportion of viruses to which a person is exposed that initiate infection. This occurs in a time-dependent manner after the beginning of treatment, and is represented in our model in terms of a modification to the exposure distribution (Figure 1C). Our model describes a situation in which UDCA treatment reduces the probability that an exposure will lead to infection, lowering the odds ratio of infection (Figure 1D).

**Figure 1:**
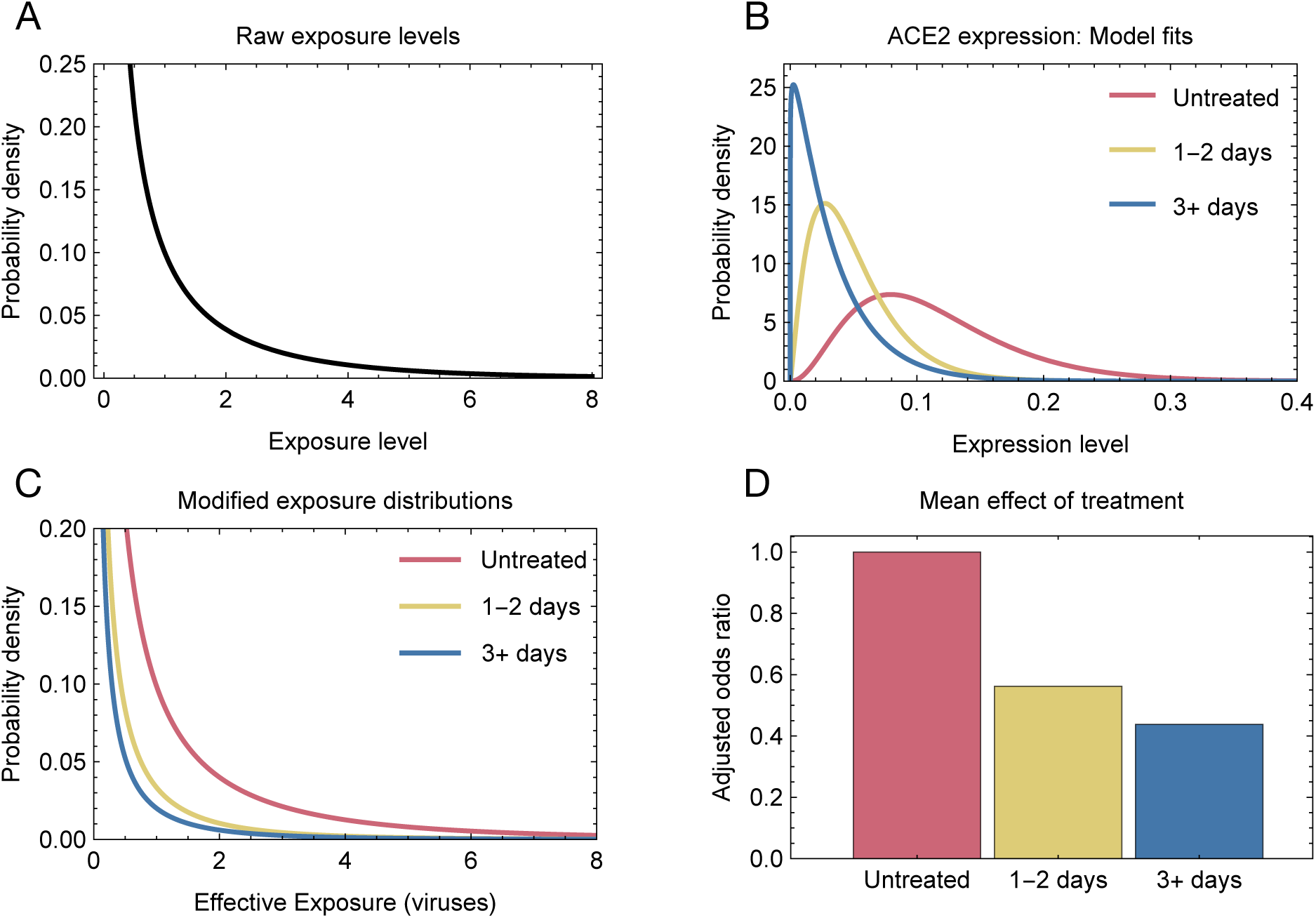
Basic model details. **A.** Distribution of SARS-CoV-2 exposure used in our model. An exposure of 1 means that an individual is infected by a Poisson random variable with parameter 1. If this random variable is equal to or greater than 1, the individual is infected, rather than not infected. **B.** Derived distribution of ACE2 expression levels in treated and untreated individuals. **C.** Effective exposure distributions for treated and untreated individuals. People who are taking UDCA are less likely to be infected by larger numbers of viruses. **D.** Adjusted odds ratios of infection calculated across a population of individuals with a distribution of ACE2 levels.

This mathematical approach enables the *in silico* evaluation of potential strategies for using UDCA to reduce SARS-Cov-2 infection; we here evaluate the potential effect of UDCA being administered to patients on a ward following the detection of a case of SARS-CoV-2. Details of the derivation of our basic model are shown in Supplementary Figures S1 to S3.

## Results

### Hospital-based transmission

We integrated our model of UDCA treatment and SARS-CoV-2 infection within a larger model describing nosocomial transmission in a hospital environment. Simulations were run to describe an environment in which UDCA was not used, with the effect of treatment being represented by retrospective changes made to each simulation. Data from an outbreak extracted from one simulation shows how UDCA had the potential to cut short chains of transmission events (Figure 2A). In this case, an infected individual, with ID number 953, was introduced onto a ward, leading to a large outbreak involving both patients and healthcare workers. We simulated an intervention in which, upon the detection of a case of SARS-CoV-2 on a ward, all patients on that ward, plus any arriving onto that ward during the intervention, were treated for 10 days with UDCA. Changes in individual probabilities of being infected upon the intervention are shown in Figure 2B. Here, the ward onto which patient 953 was introduced was already marked as an intervention ward, so that patients in contact with patient 953, and those further down the network, were already treated with UDCA. In our simulation this reduced the probability of those patients being infected, further reducing the probabilities of individuals further down the transmission chain from being infected. Healthcare workers in the network were not treated, but acquired a lower risk of infection via a reduction in exposure to infected patients. Across multiple transmissions within the network the probability of an individual being infected can be greatly reduced; for example the intervention results in the individual 4665 having only a 7.5% chance of being infected. Within this network the numbers of secondary infections were reduced from 7 patients and 8 HCWs to a mean of 1.9 patients and 4.6 HCWs (Figure 2C/D).

**Figure 2:**
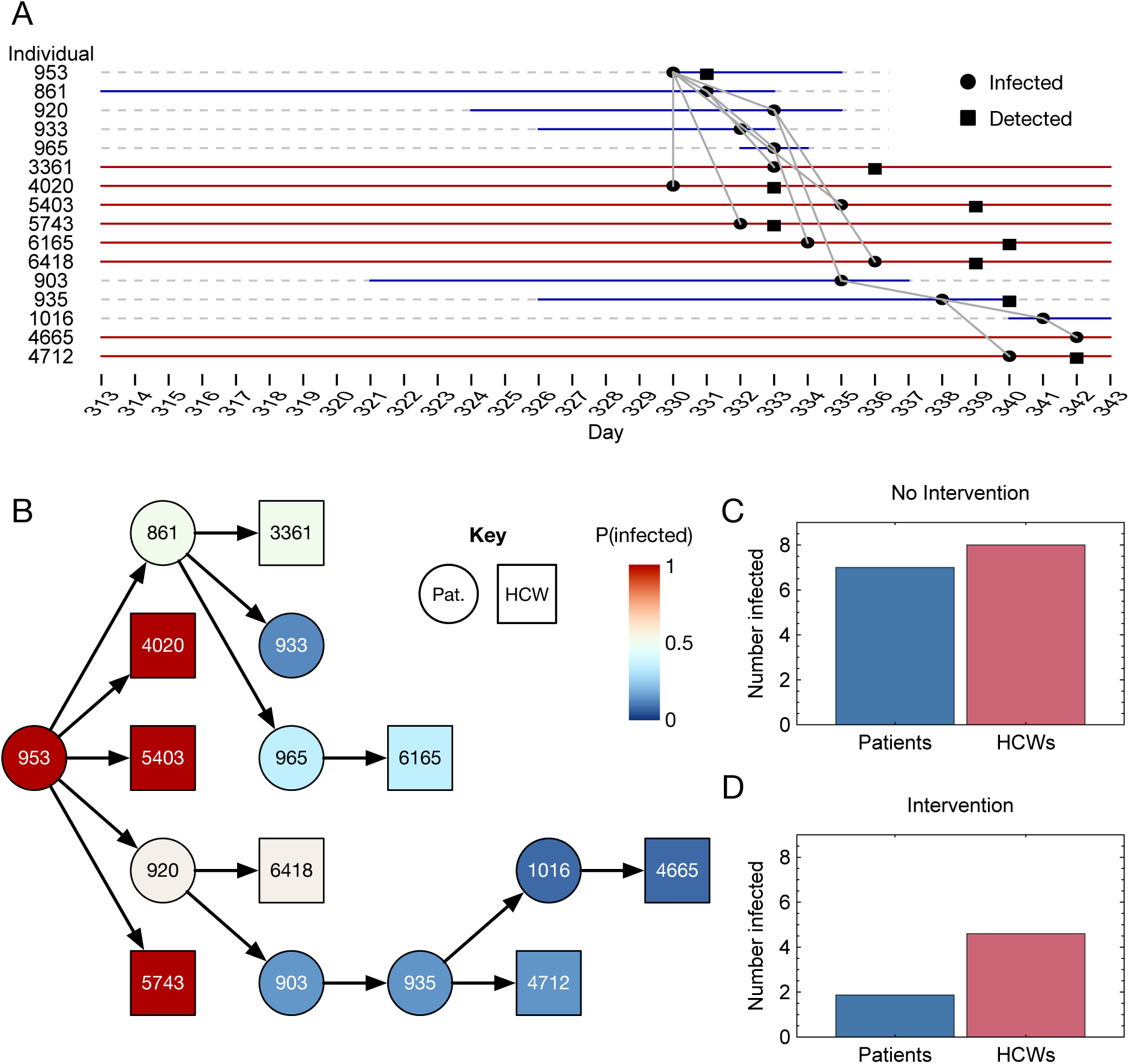
Application of UDCA treatment to a transmission network. **A.** Case of nosocomial transmission within a hospital simulation. Horizonal lines show times when individuals were in the hospital, coloured blue for patients or red for HCWs. Black dots indicate dates on which individuals were infected, while squares indicate dates on which cases of infection were detected, where detection occurred. **B.** Representation of one realisation of the impact of UDCA treatment upon the transmission network. The ward upon which the outbreak began was an intervention ward at the time of patient 953 being introduced onto the ward. UDCA treatment reduced the probabilities of patients being infected with SARS-CoV-2, and by implication reducing the probabilities of their further transmitting the virus. The reductions in individual probabilities caused by UDCA are stochastic, representing random samplings of the baseline ACE2 levels of expression of individuals. **C.** Numbers of secondary cases of SARS-CoV-2 infection in patients and HCWs in the network in the absence of an intervention. **D.** Numbers of secondary cases given treatment with UDCA.

Applied to a set of simulated data, describing nosocomial transmission in 60 hospitals over a window of 610 days, our simulated intervention reduced cases of nosocomial transmission among patients by 16.5% (95% C. I. 14% - 20%) (Figure 3A). As modelled, the intervention reduced nosocomial infection of HCWs by close to 4% (Figure 3B). These figures are lower than obtained for the example network of Figure 2, which was an outlier in terms of its large size. Across hospital simulations, the most common instance of nosocomial transmission involved one person infecting another, with no further transmission. The mean transmission network involved 3.1 individuals, or equivalently 2.1 cases of nosocomial transmission (Supplementary Figure 3).

**Figure 3.**
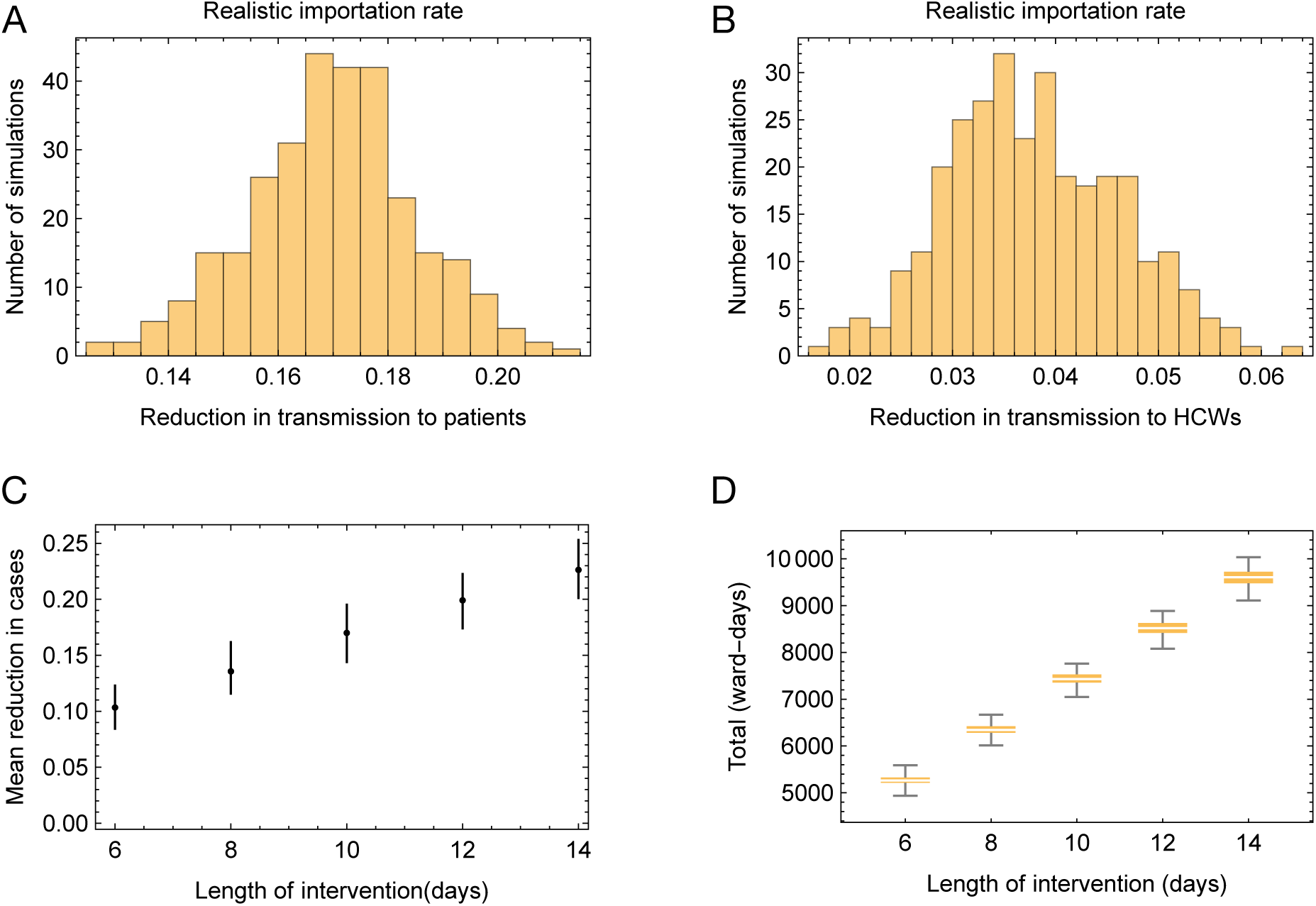
Reduction in transmission caused by the simulated use of UDCA. **A.** Reductions in nosocomial transmissions to patients, measured across five stochastic replicates of each of 60 hospital simulations. **B.** Reductions in nosocomial transmission events to healthcare workers, measured across five stochastic replicates of each of 60 hospital simulations. **C.** Variation in the reduction in nosocomial transmissions to patients, varying the length of the intervention, given the detection of a case upon a ward. **D.** Total length of interventions in a hospital according to the length of an individual intervention, measured in ward-days, across the 624 days of the simulation.

Our results were sensitive to the parameters of our model. For example, changing the window of intervention following the detection of a case led to changes in the reduction of nosocomial transmission among patients, from 16.5% at the default 10-day window of intervention to 10.3% for a 6-day window of intervention, or to 22.6% for a 14-day window of intervention (Figure 3C). Changing the length of the intervention window led to proportional changes in the number of ward-days for which the intervention was in place. Where under default parameters, the intervention was in place for a mean of 7440 ward-days, this statistic ranged between 5270 ward-days given a 6-day intervention to 9610 ward-days given a 14-day intervention (Figure 3D).

The effect of the intervention was also sensitive to the simulated level of community infection, which in our model determines the rate of importation of SARS-CoV-2 patients into a hospital. Halving the importation rate compared to the default led to an estimated 11% reduction in nosocomial cases in patients, while doubling the rate of importation relative to default led to an estimated 24% reduction in nosocomial cases (Figure 4A, B). Although a higher importation rate led to more cases of SARS-CoV-2 in hospital, and an increased number of cases of nosocomial transmission, the distribution of the sizes of transmission networks was not substantially altered (Figure S4). Rather, the difference in outcome arises from an indirect effect of the number of cases: A greater number of cases in hospital meant that more transmission networks occurred on wards that had previously been marked out for intervention. UDCA has a time-dependent effect on ACE2 expression levels. In a hospital where SARS-CoV-2 outbreaks are sparse, there are few wards on which the intervention is in place. Given a new outbreak there is a delay arising from the time taken to detect a new outbreak, and then the time taken for UDCA treatment to have full effect, before the full impact of the drug is experienced. By contrast, where outbreaks are common, the probability of a new outbreak arising on a ward in which the intervention is already in place is increased. In such a case the delay is reduced, with UDCA potentially providing its maximal benefit from the first day of the outbreak. We note that, at the modelled high rate of importation, a situation is reached in which, at peak, nearly all hospital wards are included in the intervention (Figure 4C, D). At this point any new cases of nosocomial transmission occurring in the hospital involve already-treated patients, maximising the effect of the intervention.

**Figure 4:**
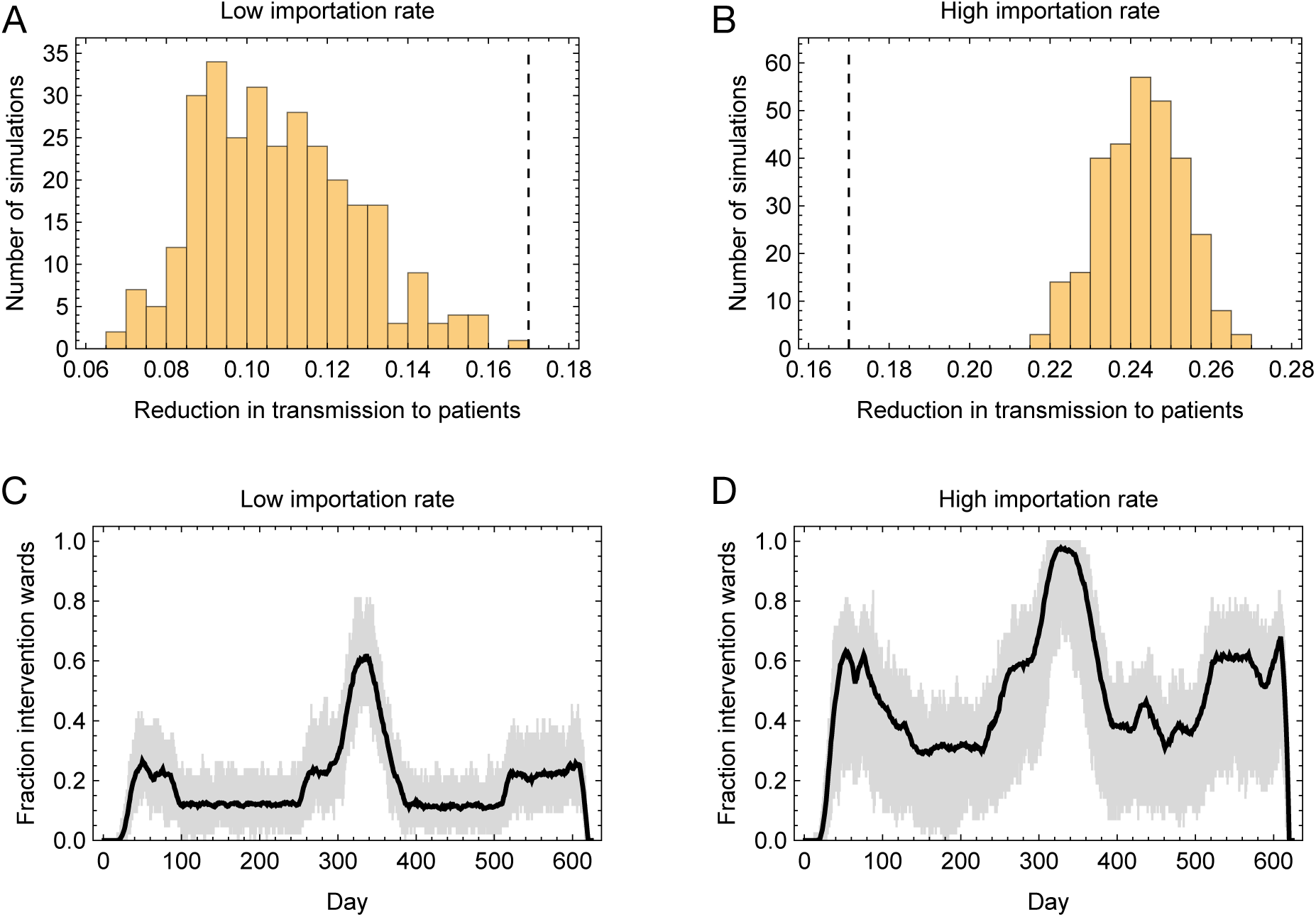
Effect of variation in the rate of importation of SARS-CoV-2 cases to the hospital. **A.** Reductions in nosocomial transmissions to patients, calculated under a low importation rate. The vertical black dashed line shows the mean of this statistic under a realistic importation rate, modelling importations in the UK between 2020 and 2021. **B.** Reductions in nosocomial transmissions to patients, calculated under a low importation rate. The vertical black dashed line shows the mean of this statistic under a realistic importation rate. **C.** Fraction of wards in which the intervention was taking place, plotted by time. The black line shows the mean calculated across 60 simulations with a low importation rate. The gray shaded region shows the range across simulations. **D.** Fraction of wards in which the intervention was taking place, plotted by time. The black line shows the mean calculated across 60 simulations with a high importation rate. The gray shaded region shows the range across simulations.

## Discussion

We have here considered the use of a potential candidate for use as pre-exposure prophylaxis against SARS-CoV-2 infection. Our approach combines mathematical models of changes in ACE2 expression, the consequences of those changes for virus entry into cells, and the likely levels of exposure to the virus encountered by individuals; combined these models produce a result consistent with observational studies. Applying our model to simulated data describing a hospital setting, our approach suggests that a proposed intervention using the drug might lead to a reduction of around 17% in nosocomial transmission to patients. Our work provides an indicative and preliminary assessment of the potential for UDCA as to be used for pre-exposure prophylaxis to combat nosocomial transmission.

Our modelling provides some insight into the potential value of pre-exposure prophylaxis in a real-world setting. Where modelling has suggested the use of pre-exposure prophylaxis as an effective tool to prevent SARS-CoV-2 infection, the implicit need for action to be taken prior to an exposure limits its value. In modelling a hospital outbreak we have chosen a setting in which the potential risk of infection is unusually high, in which cases of infection are being actively monitored, and in which treatment could be administered by competent medical professionals. In this setting, the time-dependent effect of treatment was predicted to have an effect. A proportion of the effectiveness of the intervention is explained by the reduction in transmission events withing outbreaks that occur on an intervention ward but are unrelated to the outbreak which triggered the intervention.

In this sense, our work highlights a distinction between preparatory and responsive approaches to reducing infectious disease transmission. Preparatory approaches, such as improving ventilation, mask-wearing, and hand hygiene, create an environment in which transmission is intrinsically less likely. Responsive approaches, which are implemented following the detection of an outbreak, are limited by imperfect surveillance, and in the case of UDCA, by transmission events having occurred prior to the intervention taking effect. Our calculated reduction in transmission of 17% compares to a recent study suggesting a 22% reduction in transmission with the use of improved ventilation^17^, but may be optimistic. The levels of community importation into the hospital may be lower than reflected in the assumptions underlying the simulation, such that the effectiveness of the intervention we model would be reduced.

Our model has some implications regarding SARS-CoV-2 transmission, for example that people with higher baseline levels of ACE2 expression are more likely to be infected with SARS-CoV-2. Where someone’s baseline ACE2 expression level is high, more of the viruses they are exposed to are likely to gain entry into cells and initiate infection. Our model also implies that a high baseline ACE2 expression level will reduce the efficacy of UDCA in preventing infection. UDCA reduces the number of viruses entering cells, but to prevent infection this number must be reduced to zero. The reduction to zero is less likely when the initial number, due to high ACE2 expression, is large. Figure S5 describes expected reductions in transmission for high and low baseline levels of ACE2 expression.

Our modelling approach is built upon multiple assumptions. For example, our model of exposure to SARS-Cov-2 was constructed using data describing transmission in a domestic, rather than a hospital setting. The simulated hospital environment, while representing the state of the art, also contains multiple simplifications. For example, a healthcare worker who avoided being infected due to the indirect effect of UDCA would retain a risk of being infected at some later date via the local community. Our simulated approach neglected this risk: Once prevented from being infected on one occasion, HCWs stayed uninfected for the remainder of the simulation. As such our result about preventing infection in healthcare workers is open to some uncertainty.

Our model assumes that the distribution of ACE2 levels in the hospital population mirror those of the general population. Further, simulations of hospitals simplify elements such as the movement of patients and staff between wards which likely impact upon transmission. Differences between hospital environments, such as those of hospital architecture, are not captured by our model. However our simple mathematical model describing the effect of UDCA upon SARS-CoV-2 transmission gave a result consistent with that described in a cohort study. Previously, John et al found an adjusted odds ratio for risk of symptomatic COVID infection of 0.54 (confidence interval 0.39 – 0.73) among individuals with cirrhosis who were treated with UDCA^23^. Considering an individual who had been treated for at least 3 days prior to exposure to SARS-CoV-2, our model suggested that continuous treatment with UDCA would reduce the incidence of COVID infection by 56%, equivalent to an adjusted odds ratio of 0.44.

The intervention we propose is described in a simplified fashion. Although long-term therapy with UDCA is well-tolerated^34^, aspects such as negative drug-drug interactions may prevent the giving of the drug in a universal manner to a cohort of patients. We have here considered the use of UDCA for pre-exposure prophylaxis in a situation where full clinical supervision is possible. A realistic intervention might use the drug only in subsets of cases where SARS-CoV-2 infection would have particularly severe consequences, such as in a care home or with more vulnerable patients. The authors do not recommend the use of UDCA in any situation except under the explicit guidance of a qualified physician.

## Methods

We built a model to estimate the effect of treatment with UDCA upon SARS-CoV-2 transmission. Our model has three parts, the first modelling the effect of UDCA upon ACE2 expression in an individual, the second modelling the effect of changing ACE2 expression upon virus entry into cells, and the third modelling the likely distribution of exposure to SARS-CoV-2 viruses, given knowledge of transmission bottlenecks from cases of SARS-CoV-2 infection.

### Changes in ACE2 expression given treatment with UDCA

Given data from a set of individuals describing variation in ACE2 levels after commencing treatment with UDCA^23^, we used a time-dependent gamma distribution to represent the effects of the drug. In this equation x describes ACE2 expression level. The Mathematica software package was used to identify optimal parameters a and b within this model.

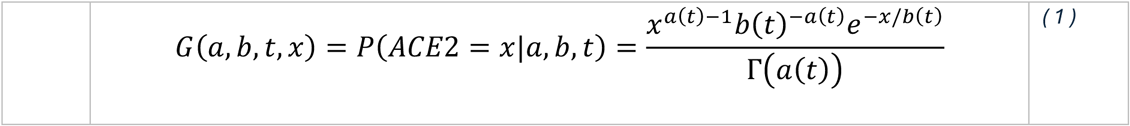

G(a,b,t) was here characterised using data describing three discrete time intervals, for t=0 days, t ∈ {1, 2} days, and t ≥ 3 days. Raw data are shown in Figure S1; the inferred distributions are shown in Figure 1B.

### Modelling of the relationship between ACE2 expression and viral infection

Data describing ACE2 expression and SARS-CoV-2 infection levels from different cell types within a lung explant was used for modelling purposes^23^. Under a model of receptor association, the relationship between ACE2 receptor availability and virus entry are expected to follow the Hill equation^35^. To explore whether, at the physiological concentrations of ACE2 represented by our data, a linear model would be a reasonable approximation to this, we fitted both linear and sigmoidal curves to the data. Where the level of ACE2 expression is given by [ACE2], our models were

Linear model:

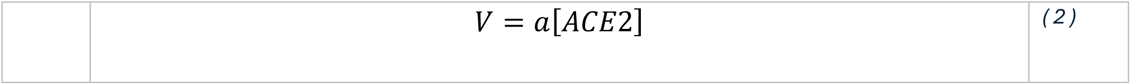

Hill equation:

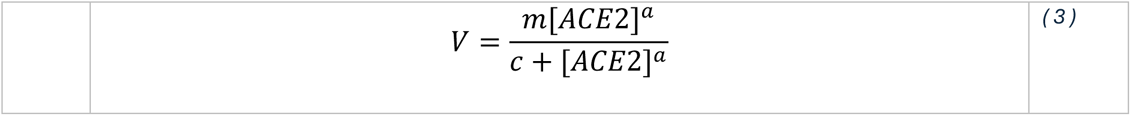

Models were fitted to the data using the NonlinearModelFit routine within the Mathematica software package, inferring different parameters for the lung and bronchaeal data; these data were not directly comparable due to different cell compositions of the different samples. Model fits were compared using the Bayesian Information Criterion (BIC)^36^. The difference between the BIC values for the models, of 2.81 units in favour of the Hill equation model, was interpreted as positive support for this model but not strong support, following Raferty et al^37^. Data used in this modelling, and model fits, are shown in Figure S2.

### Inference of a distribution of levels of exposure to virus

We generated a model of the number of viruses expected to initiate infection during SARS-CoV-2 transmission. Given a level of exposure to viruses E, we modelled the transmission bottleneck size N_b_ (i.e. the number of viruses initiating an infection^38^) as being Poisson distributed with parameter E.

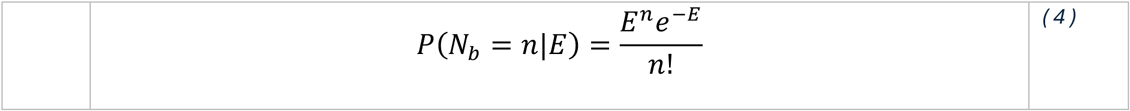

We assumed that exposure can be described by a Gamma distribution, such that E obeys the formula

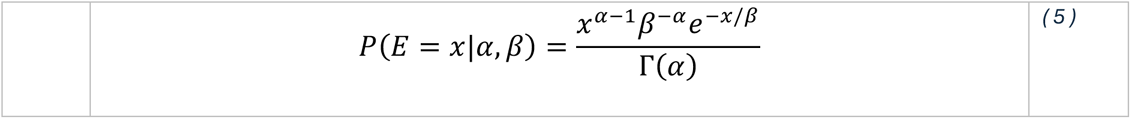

Where Γ(α) describes the gamma function

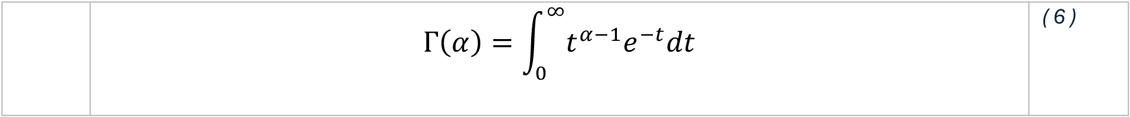

We next inferred parameters α and β for this model using data from two publications describing household SARS-CoV-2 transmission. A study of SARS-CoV-2 transmission in households inferred bottleneck sizes for 20 cases of infection^32^. We took these sizes as being indicative of viral transmission events. Next, we used the result from a meta-analysis of household transmission studies, of a secondary attack rate for SARS-CoV-2 of 18.9%^33^, to estimate that for each 20 cases of infection, there were an additional 83 cases of exposure not leading to infection. We thus compiled a dataset of 103 outcomes of exposure, 20 of which matched the bottlenecks inferred by Lythgoe et al, and 83 of which involved zero viruses. Data describing inferred bottlenecks for these 20 cases is shown in Figure S3.

For a given set of parameters α and β, we represented the gamma distribution by a discrete set of 999 equally spaced quantiles E_i_(α, β). We then calculated the mean likelihood of observing each bottleneck n_j_ in the dataset, summing these over the different bottleneck sizes.

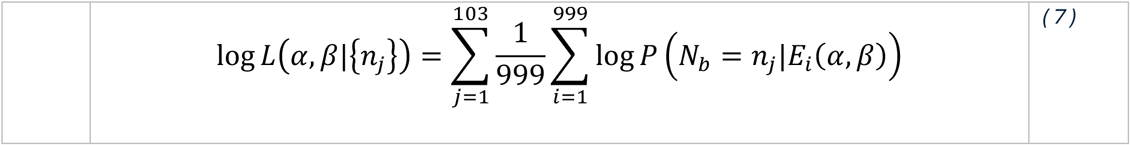

Values of α and β which maximised this likelihood were found using the Mathematica software package. We inferred the values α = 0.1558 and β = 2.797.

### Combined model of viral transmission in the presence or absence of treatment with UDCA

Under the assumption of a linear relationship between ACE2 expression and viral infection, we derived a model of viral transmission in treated and untreated individuals in a heterogeneous population. In our model, a person p receives a stochastic exposure e to viruses characterised by a random draw from the Gamma distribution E(α, β).

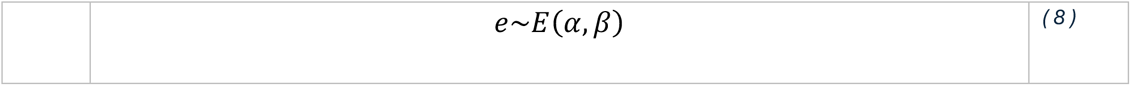

This exposure was scaled in a linear fashion by the individual-specific term I(r, t), which describes the relative ACE2 expression of the person. Here the parameter r was uniformly sampled from the interval (0,1); for example r=0.12 would imply that I(r, t)=x was equal to the 12^th^ centile of the distribution G(a, b, t, x). In this manner, the effective exposure of a person p who had undergone t days of treatment with UDCA, was described as

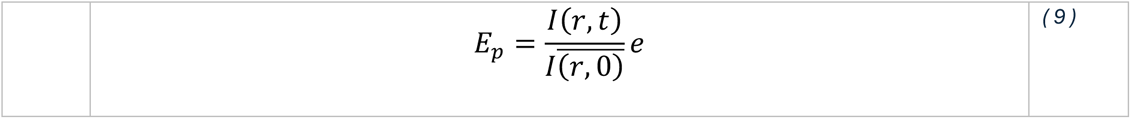

where the denominator represents the mean level of ACE2 expression for an untreated individual. The number of viruses initiating infection in this person was then given by the Poisson random variable

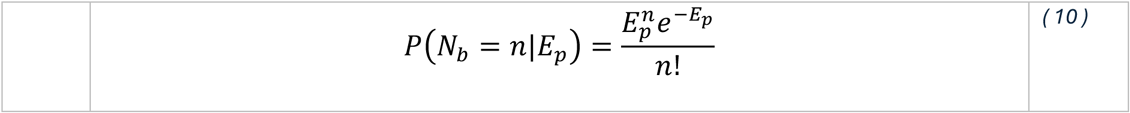

In this model, infection occurred if N_b_ was greater or equal to 1, and did not occur if N_b_ was equal to zero. Basic statistics of infection were calculated across distributions of ACE2 expression for treated and untreated individuals, integrating over a representative range of levels of exposure.

### Simulation of hospital transmission events

We used a published individual-based model of nosocomial transmission to simulate SARS-CoV-2 in hospitals, including the importation of cases from the community, the occasional spread of infection between patients and health care workers (HCWs) in hospitals, and the detection or non-detection of these cases^39^. Within this modelling framework simulations were conducted describing high, realistic, and low levels of importation of cases into hospitals. ‘Realistic’ levels were derived from now-casted community levels of infection for the East of England, spanning a window of 600 days during the SARS-CoV-2 pandemic^40^. High and low levels represented 2-fold and 0.5 fold changes to these values. For each level of importation, twenty sets of parameters representing rates of transmission to and from healthcare workers and patients were generated, consistent with evidence from a study of transmission in hospitals^41^. Three statistical replicates were generated for each set of parameters, making a total of 180 simulations.

For each simulation, transmission networks were identified, comprising sets of patients and HCWs who infected each other in hospital, alongside the dates of these transmission events. We modelled an intervention in which, upon the detection of a case of SARS-CoV-2, all patients on the ward in which the case was detected were given a 10-day course of treatment with UDCA. Where new patients arrived on the ward within 10 days of the commencement of the intervention, these patients were also started on a 10-day course of UDCA treatment. We then retrospectively altered the identified transmission networks. Transmission events in these networks were assigned a probability of occurrence equal to one. These probabilities were then altered according to UDCA treatment, with a treated patient having a reduced probability of being infected. Alterations in probabilities were then evaluated in a compound manner. For example, if in a simulation person A infected person B, who infected person C, treating person B would reduce the probability of person C being infected, even if C was untreated. The expected reduction in the number of cases of SARS-CoV-2 nosocomial infection was therefore calculated.

Our model of nosocomial transmission accounted for the variation in baseline susceptibility to SARS-CoV-2 infection implied by our model of infection and treatment. To achieve this we generated a distribution of the baseline ACE2 expression of an individual conditional upon their having been infected with SARS-CoV-2. This distribution differs from the gamma distribution fitted to the untreated ACE2 expression levels: The fact of having been infected increases the probability of an individual having a high level of baseline ACE2 expression.

To calculate the conditional distribution of ACE2 we calculated quantiles of the distributions of baseline ACE2 expression G(a, b, 0, x), and of the exposure distribution E(α, β)(y), calculating the products of these values. We then identified probabilities of infection for each datapoint, normalising these to sum to one

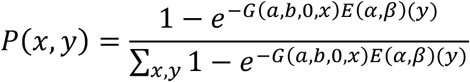

These values were then normalised to sum to one across all x and y, before summing across exposures y to calculate a probability distribution from which values of ACE2 expression could be sampled. In evaluating nosocomial transmission, individuals who were infected in the initial simulation were assigned baseline levels of ACE2 expression from this discrete distribution before evaluating the individual-specific effect of UDCA upon their probability of having been infected. The discrete distribution and the inferred probability density function are shown in Supplementary Figure S6.

## Availability of code

Code used for processing simulation data is available at https://github.com/cjri/HospitalSimulationAnalysis

## Data Availability

All data produced in the present study are available upon reasonable request to the authors. A full dataset will be made available online upon the publication of the manuscript in a peer-reviewed journal.

## Acknowledgements

This research is jointly funded by the UK Medical Research Council (MRC) (MC_UU_00034/6) and the Foreign Commonwealth and Development Office (FCDO) under the MRC/FCDO Concordat agreement.

## Supplementary Figures

**Figure S1.**
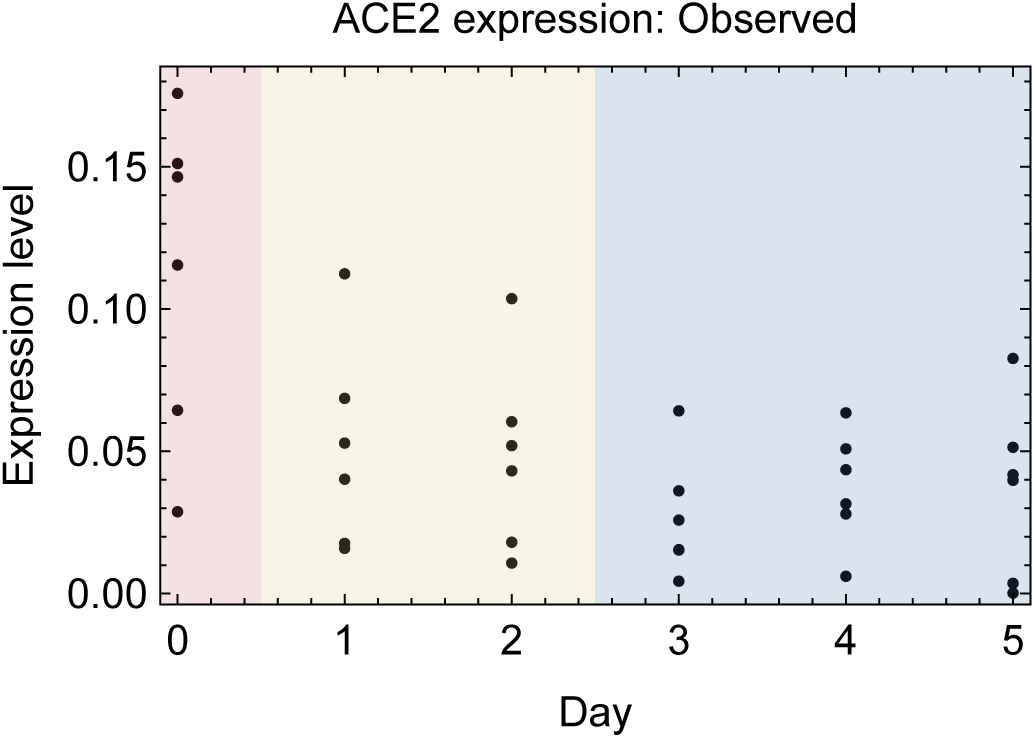
Data describing changes in ACE2 expression following the 0use of UDCA. qPCR measurements of the levels of ACE2 in nasal epithelial cells collected with nasopharyngeal swabs from six individuals who received 15 mg per kg per day of UDCA for five days, previously described by Brevini et al^23^. Shading indicates windows of time into which samples were divided for model fitting. A gamma distribution was fitted to the data within each window.

**Figure S2:**
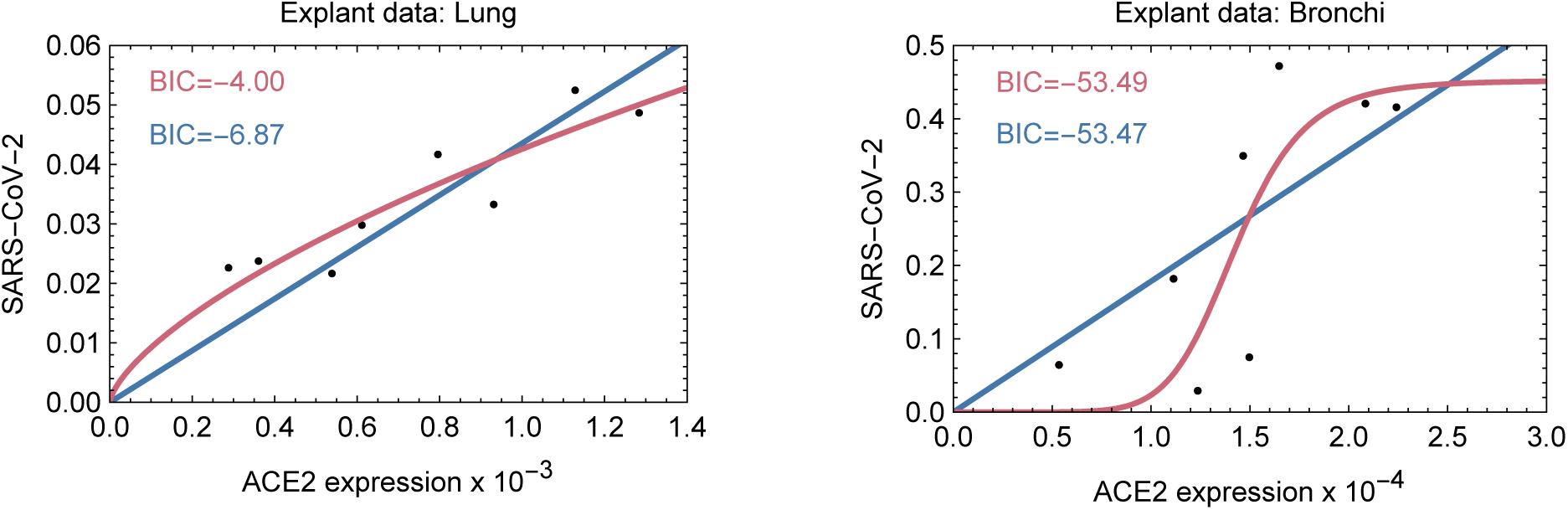
Model fits to qPCR data describing levels of ACE2 expression and SARS-CoV-2 infection in a human explant. Data describing samples collected from the lung and bronchi were originally described by Brevini et al^23^. Model fits describe sigmoidal (red) and linear (blue) relationships between ACE2 and SARS-CoV-2 expression values. Under the Bayesian Information Criterion, there was positive evidence, though not strong evidence, favouring the sigmoidal model, which follows the Hill equation. For the purpose of modelling a simple linear approximation was made.

**Figure S3:**
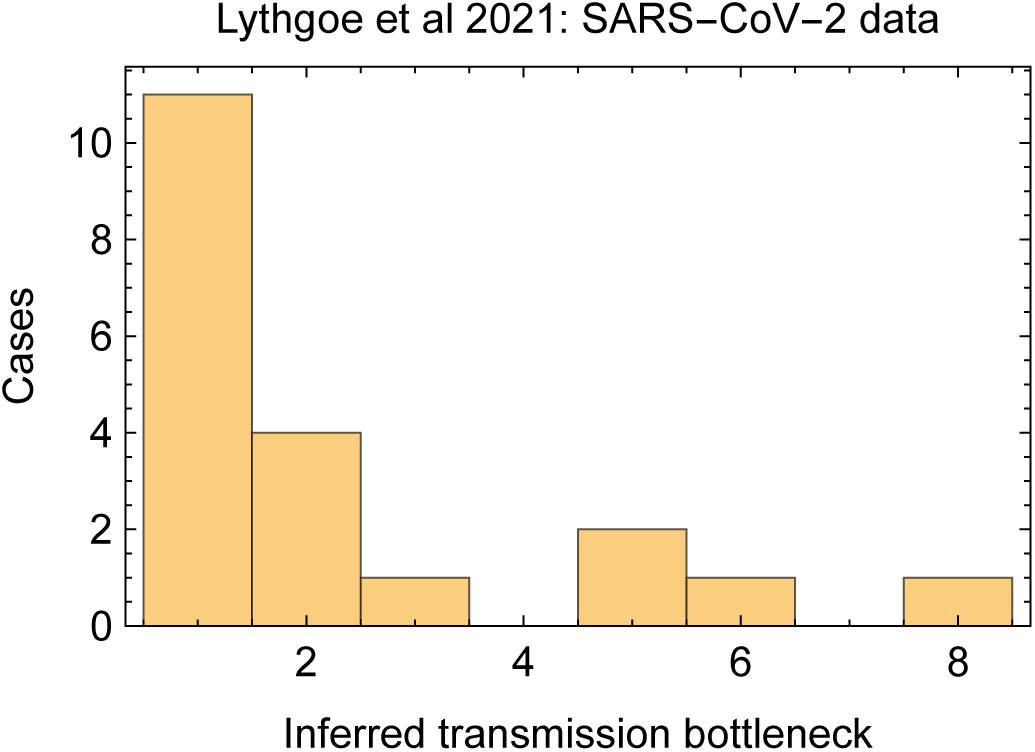
Data describing SARS-CoV-2 transmission bottlenecks, from an earlier publication^32^. Data describe the number of SARS-CoV-2 viruses initiating infection in 20 cases of household transmission. In our study these data were augmented with cases describing non-infection, that is with transmission bottleneck zero, reflecting a published secondary attack rate for SARS-CoV-2 in a domestic context^33^. Our basic exposure model, described in Figure 1A, was then fitted to the augmented data.

**Figure S4:**
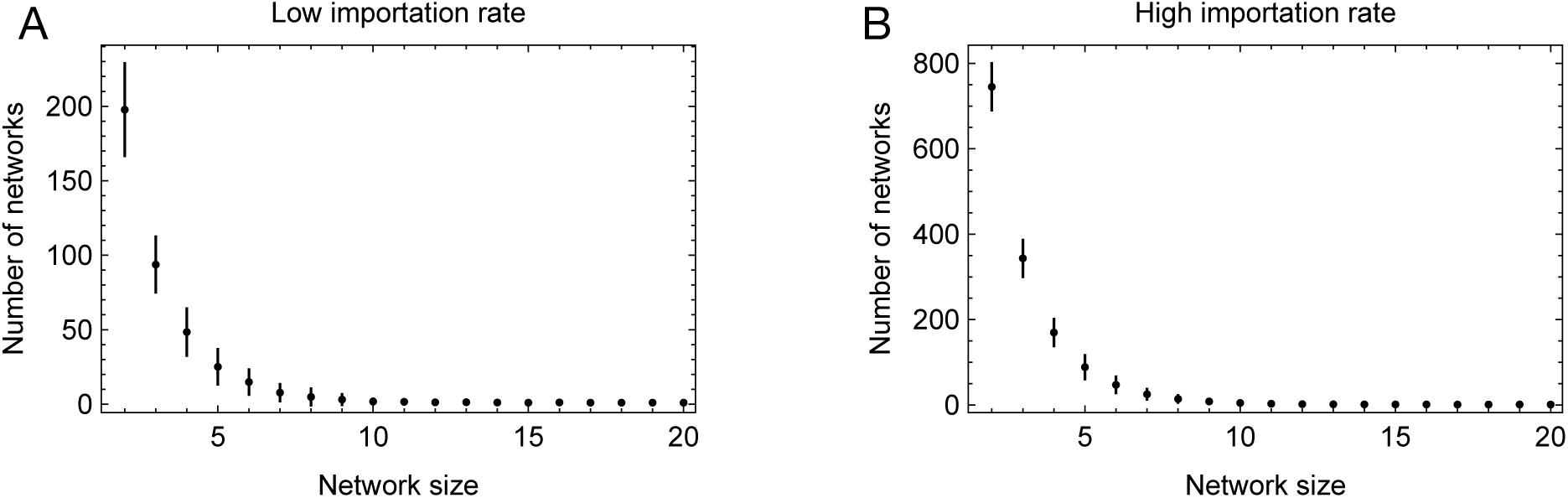
Network size numbers and distribution for different simulated importation rates. A cluster size of 2 indicates one person infecting another, with no further transmission.

**Figure S5:**
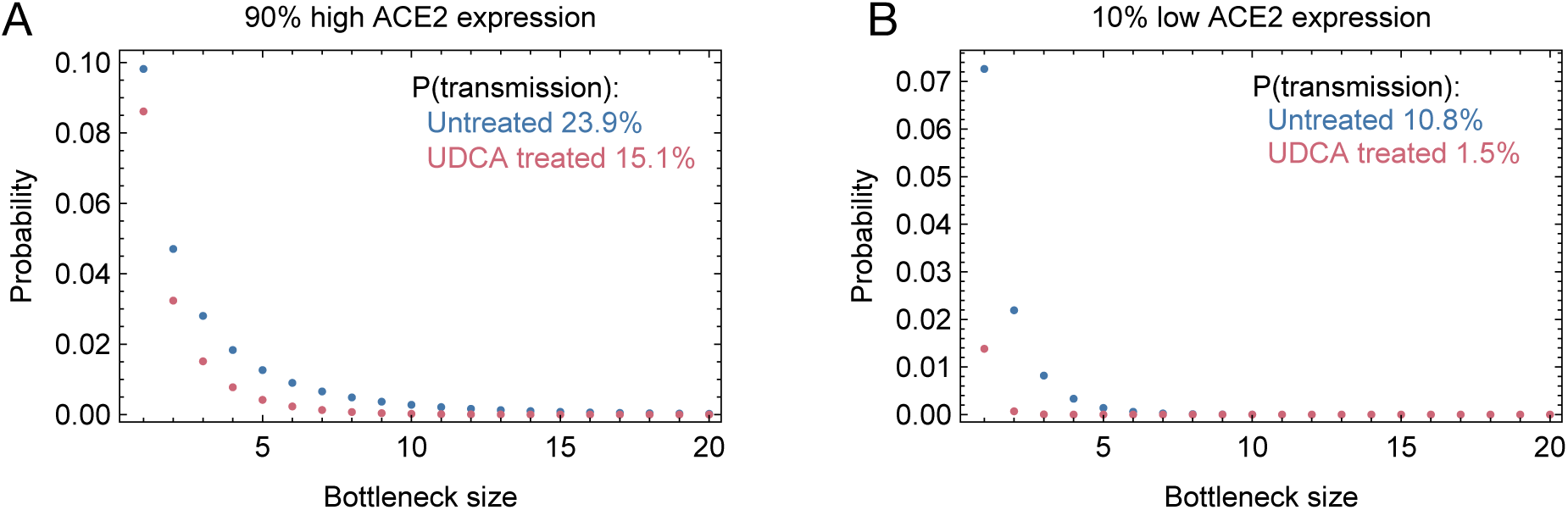
Expected distributions of SARS-CoV-2 transmission bottlenecks. Inferred values are shown for among individuals at the 10^th^ and 90^th^ centiles of ACE2 expression, in untreated individuals, and in individuals treated for more than three days with UDCA. In both cases UDCA reduces expected bottleneck sizes, but with a more dramatic reduction in the probability of transmission (i.e. bottleneck size ≥ 1) among individuals with low ACE2 expression.

**Figure S6:**
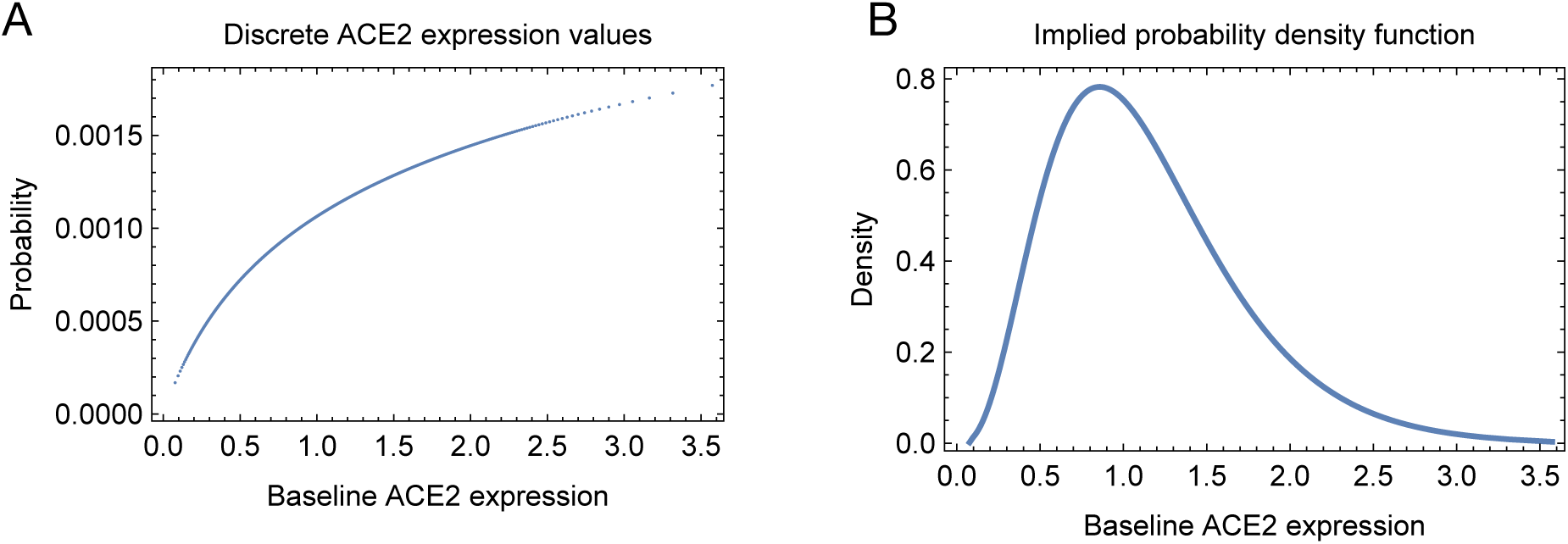
Distribution of baseline ACE2 expression conditional upon an individual having been infected. **A.** Probabilities of discrete values of baseline ACE2 expression. **B.** Probability density function implied by the discrete distribution.

## Notes

### Competing Interest Statement

The authors have declared no competing interest.

